# Epidemiology, pathways, patterns of care and Day-14 mortality of severe cases according to hypoxemia among IMCI children using routine Pulse Oximeter decentralized at Primary Healthcare in West Africa: the AIRE cohort study in Burkina Faso, Guinea, Mali and Niger, 2021 - 2022

**DOI:** 10.1101/2024.10.18.24315758

**Authors:** Hedible Gildas Boris, Sawadogo Abdoul Guaniyi, Zair Zineb, Kargougou G Désiré, Méda Bertrand, Peters-Bokol Lucie, Kolié Jacques S, Louart Sarah, Ouédraogo Yugbaré Solange, Diakite Abdoul Aziz, Diallo Ibrahima Sory, Abarry Souleymane Hannatou, Busière Sandrine, Lamontagne Franck, Shepherd Susan, Ridde Valéry, Leroy Valériane, the AIRE Research Study Group

## Abstract

**Background:** The AIRE project has implemented routine Pulse Oximeter (PO) use in Integrated Management of Childhood Illness (IMCI) consultations to improve the diagnosis and care management of severe illnesses in primary health centre (PHC) in Burkina Faso, Guinea, Mali and Niger. We analysed care management of severe cases according to hypoxemia, and the determinants of their Day-14 mortality.

**Methods:** All children under-5 attending IMCI consultations using PO and classified as severe cases (severe IMCI cases or with severe hypoxemia: SpO2<90%) were enrolled at 16 research PHCs (four/country) in a 14-Day prospective cohort with parental consent. Care management according to hypoxemia severity and determinants of Day-14 mortality were analysed.

**Results:** From June 2021 to June 2022, 1,998 severe cases, including 212 (10.6%) aged <2 months were enrolled. Severe hypoxemia was common (7.1%), affecting both respiratory cases (9.9%) and non-respiratory cases (3.7%); 10.5% had moderate hypoxemia (90%≤SpO2≤93%). Overall, 463 (23.2%) have been hospitalised. At Day-14, 95 (4.8%) have died, and 27 (1.4%) were lost-to-follow-up. The proportions of referral decision, hospitalisation and oxygen therapy were significantly higher for severe hypoxemic cases (83.8%, 82.3%, 34.5%, respectively) than for those with moderate hypoxemia (32.7%, 26.5%, 7.1%, respectively) or without hypoxemia (26.3%, 17.5%, 1.4%, respectively). Similarly, Day-14 mortality rates were 26.1%, 7.5% and 2.3% respectively (p<0.001). Death occurred within a median delay of one day for severe hypoxemia. In an adjusted mixed-effect Cox model, age <2 months, severe and moderate hypoxemia, severe malaria, and place of case management elsewhere than at PHC independently increased mortality at Day-14.

**Conclusion:** Both severe and moderate hypoxemia were frequent among outpatient critically ill children diagnosed using PO, and associated with a high mortality. Although, the diagnosis of hypoxemia prompted their care management, hospital referral and access to oxygen remain sub-optimal and crucial levers for reducing under-5 mortality in West Africa.

**Study registration number:** PACTR202206525204526 Registered on 06/15/2022

**What is already known on this topic?:**

- Under-5 mortality is high and severe hypoxemia is a strong predictor of death reported in East African studies conducted in primary care level among severely ill children
- Hypoxemia is underdiagnosed clinically leading to delayed referral and access to oxygen therapy.
- Pulse Oximetry is a simple, low cost and reliable tool to diagnose hypoxemia at a decentralized level.
- Few studies have explored the pathways and patterns of care of severe cases identified at primary care level using Pulse Oximetry, but none conducted in West Africa.

**What this study adds?:**

- Hypoxemia is frequent among children under-5 with serious illnesses in Burkina Faso, Guinea, Mali and Niger: 17.6% overall (severe: 7.1% SpO2<90% and moderate: 10.5% SpO2[90-93%]), higher in neonates, and affecting both respiratory and non-respiratory cases.
- Implementing systematic PO use into Integrated Management of Childhood Illness consultations has improved clinicians’ decision in case management of severe cases. It showed an increasing gradient of care management indicators according to hypoxemia: referral decision, effective hospitalization and oxygen therapy rates were significantly higher for severe hypoxemic cases compared to those with moderate hypoxemia, and those without hypoxemia.
- Nevertheless, hospitalisation and access to oxygen remain sub-optimal.
- Day-14 mortality rate was high, occurred mainly during hospital transfer or at hospital admission, and was correlated with the level of hypoxemia.

**How this study might affect research, practice or policy?:** This study supports the need to:

- Reconsider the place of place of pulse oximetry and the oxygen saturation thresholds in primary care
- Update IMCI guidelines with the routine introduction of PO use at primary care to improve the diagnosis and case management of children based on risk-stratification according to severe and moderate hypoxemia
- Strengthen the hospital referral system in West Africa to ensure that all severe cases with severe hypoxemia identified at primary care will have a chance to access to oxygen available at hospital level, or consider access to mobile oxygen at PHC.

## Introduction

Since the 1990s, mortality among children under-5 has been reduced. The global under-5 mortality rate fell from 93 per 1,000 live births in 1990 to 38 in 2019, a 59% reduction (1). Despite these advances, still 5 millions of children under-5 died worldwide in 2021, particularly in Sub-Saharan Africa, where mortality rates are the highest (2). This continent faces challenges in diagnosing child-killing pathologies particularly pneumonia (24%), diarrhoea (15%) (3) and malaria (9%), with malnutrition being the underlying causes in half of deaths of children aged between 1 and 59 months (4,5). Severe hypoxemia, defined as low haemoglobin oxygen saturation <90% at the sea level as measured by pulse oximeter (PO) (peripheral oxygen saturation, SpO2) is a life-threatening condition requiring an urgent oxygen therapy (6). It is a common sign of severity among children with both acute respiratory diseases, but also non-respiratory illnesses, and increases substantially their mortality risk (7–9).

In Sub-Saharan Africa, the health care system is weak with insufficient investment in health resources, and low access to health care services. Despite several strategies put in place, such as programs to combat targeted diseases, e.g. malaria (10–12), malnutrition (13,14), challenges in making accurate illness diagnosis still remain, particularly at the level of Primary Healthcare Centres (PHC), the first entry point to the healthcare system for sick children. There, the health staff is essentially made up of nurses with low diagnosis capacity of child illnesses. Since 1996, the World Health Organization (WHO) has developed Integrated Management of Childhood Illness (IMCI) guidelines providing a clinical syndromic approach to manage the leading causes of serious illnesses in children under-5 at the PHC level (15–17). This symptom-based algorithm aims to distinguish children who are eligible for outpatient treatment when referral is not required (treatment at home for ‘green or simple cases’ and treatment at PHC then at home for ‘yellow or moderate cases’) from those with severe illness with a high risk of death requiring urgent hospital referral (‘red or severe cases’) for an adequate management. Since the 2000s, the use of malaria rapid diagnostic tests (mRDT) was introduced into IMCI guidelines (18), which has improved the diagnosis and treatment of malaria among IMCI children (19). However, the poor clinical identification of severe hypoxemia at the PHC level contributes to residual child mortality in West Africa (20). PO is a simple, low cost and reliable tool for diagnosing hypoxemia (6). Therefore, the use of PO at PHC level for an early detection of hypoxemia for respiratory and non-respiratory cases with referral for emergency oxygen therapy at hospital level could contribute to reduce their mortality. For the first time in Africa, PO has been used at PHC level in rural Malawian children with pneumonia (21,22). Until 2020, a few field experiences of PO introduction at PHC level in Africa have been reported (8,23), but none in West Africa, which requires further investigation.

The AIRE project (Amélioration de l’Identification des détresses Respiratoires de l’Enfant) has been implemented in 2021 by a consortium of three NGOs (ALIMA, SOLTHIS, Terre des hommes) and the French Institute of Health and Medical Research (Inserm). It was aimed to improve the detection of severe hypoxemia and its care management in children under-5 years of age at PHC level by introducing the routine use of PO integrated into IMCI consultations in 202 PHCs in four West African countries: Burkina Faso, Guinea, Mali, Niger (24). An operational research component was conducted: a first cross-sectional study has reported the prevalence of severe IMCI cases using PO and its correlates at PHC level (25). The present study aimed to analyse the pathways, models of care, and mortality outcomes at Day-14 according to hypoxemia of the severe cases identified at IMCI consultations using PO and included in a prospective cohort until Day-14.

## Methods

### Study sites

The AIRE operational research study had been conducted from June 2021 to June 2022 in four countries: Burkina Faso, Guinea, Mali, Niger. The project took place in two health districts per country, covering eight district hospitals and 16 research sites (4/country) out of the 202 interventional PHCs, with approval from the national health authorities (24).

### Study design

All the children under-5 years of age attending IMCI consultations at the 16 research PHCs and classified as severe cases using PO (severe IMCI cases or with severe hypoxemia: SpO2<90%) were enrolled in a 14-Day prospective cohort.

### Study population and inclusion process

From June 14^th^ 2021 to June 20th 2022, all the neonates (0 to 59 days) and older children (2 to 59 months) attending IMCI consultations at the 16 research PHCs were screened by the site HCW using the national IMCI algorithms to classify and manage them based on their disease severity into three groups: green for simple cases (going back to home), yellow for moderate cases (observed and treated at PHCs then at home) and red for severe cases requiring urgent hospital transfer. All were eligible for PO use, except children aged 2-59 months classified as simple cases and without cough or breathing difficulties. During the consultation, the PHC-based health care worker (HCW) classified them into three groups using the IMCI classification, then used PO. Children initially classified as green or yellow IMCI cases but with SpO2 less than 90% were additional severe cases joining the red group. After the IMCI consultation, all the children eligible for PO use were enrolled with parental written informed consent: non-severe cases in a cross-sectional study reported elsewhere(25). All the severe cases as defined above were enrolled in a short prospective cohort study of 14 days. Children were not included on nights, weekends and public holidays because the research teams were not on site.

### Clinical procedures and definitions

Procedures for the IMCI consultations and for carrying out the mRDT were modified by the AIRE project. PO (Acare Technology, Taiwan; AH-M1 S0002033) was used after IMCI classification to measure SpO2 also providing heart rate. HCW had received refresher training in IMCI guidelines including measurement of SpO2 using PO with appropriate probe according to age groups. Severe hypoxemia was defined as SpO2 level <90%, moderate hypoxemia when SpO2 was from 90% to 93%, and global hypoxemia when SpO2≤93%. Normal heart rate was defined as heart rate between 100 and 160 per minute in children aged from 0 to 1 year, from 90 to 150 for children from 1 to 3 years, and from 80 to 140 for those aged between 3 to 5 years (26). Respiratory rate was only measured in children aged between 2 and 59 months according to IMCI guidelines. Fast breathing was defined as a respiratory rate ≥50 breaths per minute for children aged from 2 to 11 months, and ≥40 breaths per minute for those aged 12 to 59 months (17). We defined children as ‘respiratory cases’ when the clinician identified at least one respiratory symptom such as coughing, rapid breathing, stridor… All other vital parameters (axillary temperature, weight, height…) were measured before consultation on the triage room if available, or at the beginning of IMCI consultation by HCW. This was made using thermometer, child scale, height gauge. Fever was defined as body temperature ≥38°C using digital thermometer (26). Severe cases were eligible for hospital referral.

A separate research team, made up mainly of nurses, was dedicated to research data collection at PHC and hospital levels. They extracted data from paper consultation registers where IMCI was paper-based (Guinea, Niger, 2 PHC in Mali), and from electronic IMCI database elsewhere. At the hospital, data was extracted from the children’s medical records, and health outcomes data at Day-14 were collected by home visit or telephone call.

### Data collection

Individual data collection had been carried out over the period of enrolment via electronic case report form (REDCap® software). These included socio-demographic data, clinical data about IMCI consultation (SpO2 level, main IMCI symptom-based diagnosis blocks, treatments, decision of care management), place of case management (PHC, hospital or none of those), delay of referral, International Classification of Diseases 10 (ICD-10) for the main diagnosis retained at hospital, patterns of care (oxygen therapy), and vital status outcome at Day-14.

### Data analysis

We first described the inclusion process of children under-5 eligible for PO use during IMCI consultations in the AIRE research PHCs, and the prevalence and characteristics of all severe IMCI+PO cases enrolled, stratified by countries (distribution by age, sex, other socio-demographic variables: number of people in household, education level of household manager, income-generating activity of accompanying, distance from home to PHC, rural/urban PHC). Then, we described their clinical characteristics according to age groups (% of respiratory cases, % of severe and moderate hypoxemia, % of hospital transfer decided by HCW). Third, we described their care pathways, care management and Day-14 mortality rate (with their 95% confidence interval (95% CI) according to country and hypoxemia severity. Quantitative variables were described using means and standard deviations. Categorial data were described with proportions with their 95% confidence intervals (CI) and were compared using Pearson χ2 or Fischer exact tests.

We estimated the Kaplan Meier probability of death at Day-14 according to hypoxemia and analysed their predictors of death, using an adjusted mixed-effect Cox regression model with a random effect on PHC. Variables explored in univariate, then in a stepwise descendant approach including variables associated with a p-value <0.20 were: age, sex, literacy of accompanying person, income-generating activity, travel time from home to PHC (>30 min), consultation delay (>2 days) since the onset of disease, type of IMCI support (paper or electronic), presence of respiratory symptoms, hypoxemia, heart rate, IMCI diagnosis (severe pneumonia, severe malaria, severe acute malnutrition) and the place of case management (PHC, district hospital or back to home). The variables age and sex were forced in the full model. A two-tailed P-value of < 0.05 was considered as statistically significant. R software version 4.0.5 was used for analyses.

### Ethical aspects

The AIRE research study(24) had been approved by all four national ethics committee, by WHO ethics and research committee and by Inserm Institutional Review Board and were registered in the PanAfrican Clinical Trial Registry under the number PACTR202206525204526 on 06/15/2022.

### Patient and public involvement

This study was conducted using individual data collected with the ethical committee and MOH authorization. Patients were not involved in the analysis plan or result interpretation. Patients did not contribute to the writing or editing of this manuscript.

## Results

From June 14^th^ 2021 to June 20^th^ 2022, 39,360 children under-5 attended IMCI consultations in the 16 research PHCs (supplementary figure 1); 7,760 (19.7%) were not eligible for PO use according to criteria. Among the 31,600 (80.3%) children under-5 eligible for PO use, 15,670 (49.6%) seeking services at night or over the weekends were not offered the study. Among the 15,930 remaining who were offered the study, 33 (0.2%) families refused, and 15,897 (99.7%) were included in the study with parental consent, of whom 61 (0.4%) were excluded for missing IMCI classification or wrong inclusion. Overall, 15,836 (99.6%) were analysed, of whom 13,838 (87.4%) were classified as non-severe cases, and 1,998 (12.6%) as severe cases. All the latest were followed up until Day-14, except 27 (1.4%, 9 in Mali, 18 in Niger) who were lost to follow-up.

### Socio-demographics characteristics of severe cases by country

Socio-demographic characteristics of the 1,998 severe cases included (Table 1) were similar to those of the 15,836 children included in the study. Overall, 10.6% were aged under-2 months of age, ranging from 4.5% in Guinea to 19.0% in Burkina Faso. Female children represented 48.0% of the sample. The median number of people living in the same household as the child was 5. Child’s mothers were dead in 0.6% of cases. Children were accompanied in 97.5% by their father or mother on the day of the consultation. Overall, for 60.8% of children, the head of the household had received no school education (ranging from 48.2% in Guinea to 84.7% in Burkina Faso); 71.9% of child’s accompanying person had no income-generating activity; 72.5% of cases lived within a 30 min travel distance of the PHC attended. The median delay since the onset of the first signs to the day of IMCI consultation ranged between 2 days (in Burkina Faso and Mali) and 3 days elsewhere.

**Table 1:** Socio-demographic characteristics of the IMCI severe cases included at PHC level in the AIRE research sites, 2021 June – 2022 June (N=1,998)

| Variables |  | Burkina Faso<br>N=242 | Guinea<br>N=628 | Mali<br>N=804 | Niger<br>N=324 | Total<br>N=1,998 | p value |
| --- | --- | --- | --- | --- | --- | --- | --- |
| Age groups in months | <2 months, n (%) | 46 (19.0) | 28 (4.5) | 97 (12.1) | 41 (12.7) | 212 (10.6) | <0.001 |
|  | [2 to 23 months], n (%) | 71 (29.3) | 301 (47.9) | 279 (34.7) | 195 (60.2) | 846 (42.3) |  |
|  | [24 to 59 months], n (%) | 125 (51.7) | 299 (47.6) | 428 (53.2) | 88 (27.2) | 940 (47.1) |  |
| Sex | Female, n (%) | 105 (43.4) | 311 (49.5) | 389 (48.4) | 155 (47.8) | 960 (48.0) | 0.441 |
| Number of people living in household | Median [Q1;Q3] | 5[4;8] | 5[4;6] | 9[3;16] | 6[4;8] | 5[4;9] | <0.001* |
| Mother died | Yes n (%) | 2 (0.8) | 1 (0.2) | 4 (0.5) | 5 (1.5) | 12 (0.6) | 0.06 |
|  | No, n (%) | 240 (99.2) | 627 (99.8) | 800 (99.5) | 319 (98.5) | 1986 (99.0) |  |
| Education level of the household responsible | Never attended school, n (%) | 205 (84.7) | 303 (48.2) | 488 (60.7) | 219 (67.6) | 1215 (60.8) | <0.001 |
|  | Primary school, n (%) | 22 (9.1) | 135 (21.5) | 239 (29.7) | 54 (16.7) | 450 (22.5) |  |
|  | Secondary school, n (%) | 15 (6.2) | 153 (24.4) | 73 (9.1) | 46 (14.2) | 287 (14.4) |  |
|  | University level, n (%) | 0 (0.0) | 37 (5.9) | 4 (0.5) | 5 (1.5) | 46 (2.3) |  |
| Accompanying person of the child to the IMCI consultation | Mother, n (%) | 234 (96.7) | 602 (95.9) | 791 (98.4) | 310 (95.7) | 1937 (96.9) | <0.001 |
|  | Father, n (%) | 0 (0.0) | 4 (0.6) | 1 (0.1) | 7 (2.2) | 12 (0.6) |  |
|  | Others, n (%) | 8 (3.3) | 22 (3.5) | 12 (1.5) | 7 (2.2) | 49 (2.5) |  |
| Income-generating activity of the accompanying | Yes, n (%) | 40 (16.5) | 318 (50.6) | 168 (20.9) | 36 (11.1) | 562 (28.1) | <0.001 |
|  | No, n (%) | 202 (83.5) | 310 (49.4) | 636 (79.1) | 288 (88.9) | 1436 (71.9) |  |
| Family situation of the accompanying person | Currently married/coupled, n (%) | 240 (99.2) | 609 (97.0) | 792 (98.5) | 321 (99.1) | 1962 (98.2) | 0.178 |
|  | Divorced, widowed, single, no answer, n (%) | 2 (0.8) | 19 (3.0) | 12 (1.5) | 3 (0.9) | 36 (1.8) |  |
| Travel duration Home - PHC | Less than or equal to 30mn, n (%) | 216 (89.3) | 442 (70.4) | 572 (71.1) | 218 (67.3) | 1448 (72.5) | <0.001 |
|  | More than 30mn, n (%) | 26 (10.7) | 186 (29.6) | 232 (28.9) | 106 (32.7) | 550 (27.5) |  |
| Means to get to the PHC | Foot/Cart/Bike, n (%) | 152 (62.8) | 182 (29.0) | 106(13.1) | 192(59.2) | 632(31.6) | <0.001 |
|  | Car/Motorcycle/Bus: public or private transport, n (%) | 92(38.0) | 403(64.1) | 706(87.8) | 133(41.1) | 1334(66.8) |  |
| IMCI consultation delay since onset of signs (days) | Median [Q1;Q3] | 2[1;2.25] | 3[2;4] | 2[1;3] | 3[2;5] | 2[2;4] | <0.001* |
|  | Missing, n (%) | 34 (14.1) | 2 (0.3) | 21 (2.6) | 14 (4.3) | 71 (3.6) |  |

\*Kruskal-Wallis Test

### Hypoxemia and clinical characteristics of severe cases by age groups

Of the 1,998 severe cases, hypoxemia (SpO2≤93%) was frequent among severe cases at IMCI consultations reaching, 17.6% overall; 142 children (7.1%) were diagnosed with severe hypoxemia, affecting both respiratory cases (9.9%; 107/1,075), and non-respiratory cases (3.7%; 33/897); 211 (10.5%) had moderate hypoxemia. Among the 212 neonates (10.6% of the sample), hypoxemia reached 34.9% (15.1% severe and 19.8% moderate), while among the 1,786 children aged 2-59 months (89.4%), hypoxemia was estimated at 15.7% (6.2% severe and 9.5% moderate) (Table 2).

**Table 2:**
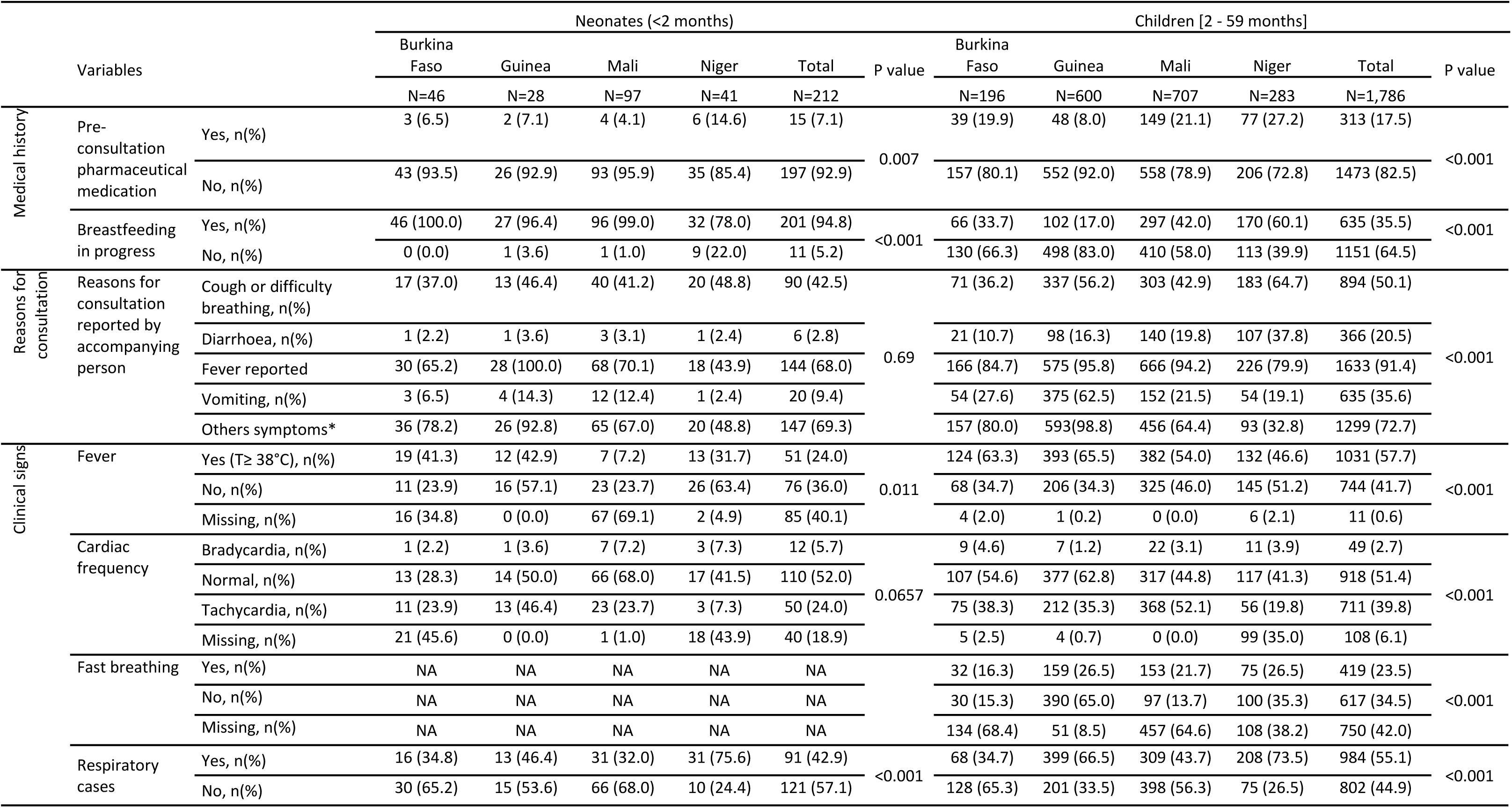
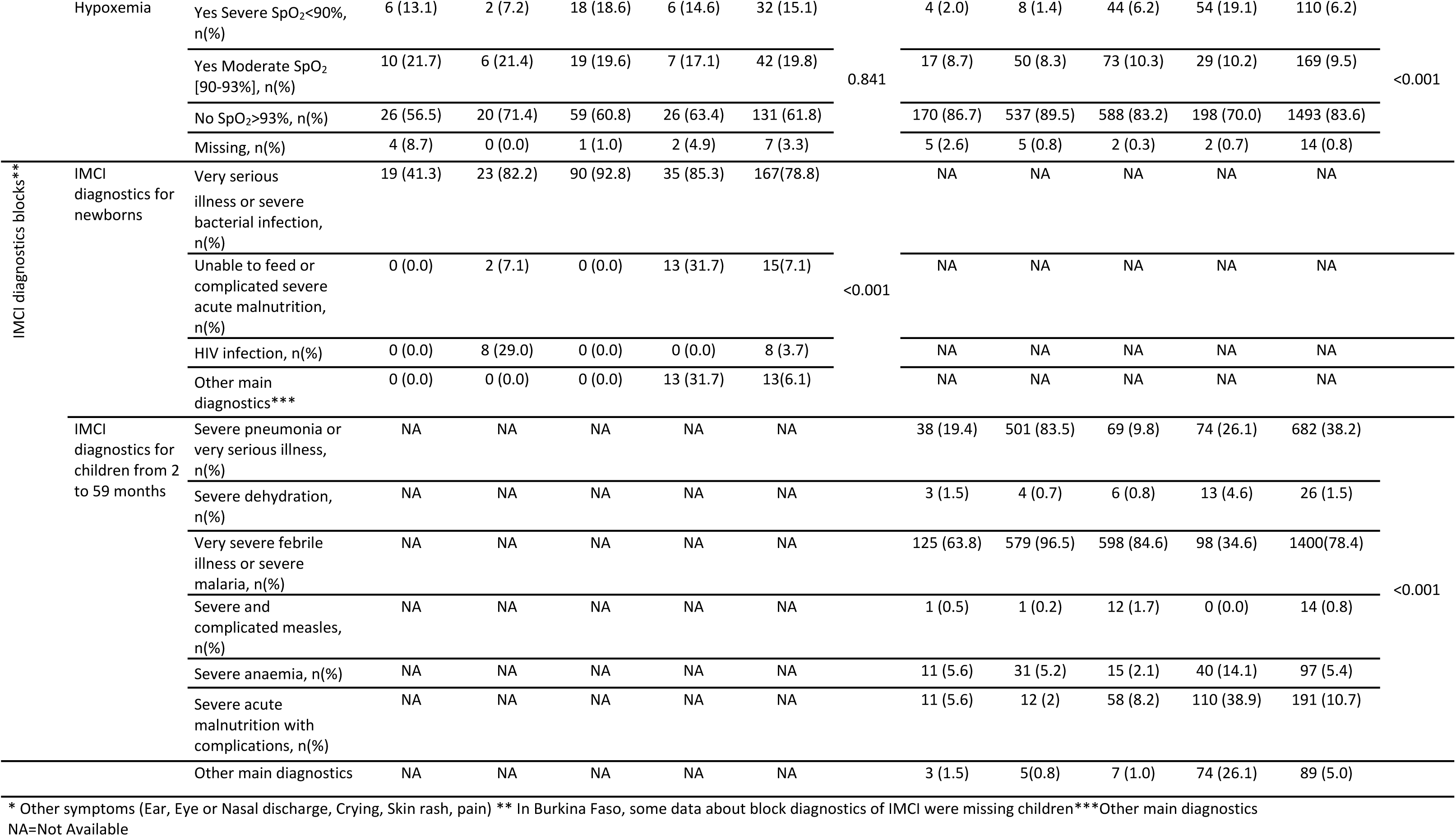
Medical history and clinical characteristics of IMCI severe cases included at PHC level in the AIRE research sites according to age, June 2021– June 2022 (N=1,998)

Among the 212 neonates, 7.1% had received pharmaceutical medication prior to the IMCI consultation. Main reasons for their consultation reported by child accompanying person were: fever (68.0%), cough or breathing difficulties (42.5%), vomiting (9.4%) and diarrhoea (2.8%). Fever was measured by the clinician in 38.7%. Fifty neonates (50.0%) had tachycardia at the time of the IMCI consultation, and 42.9% had respiratory signs. Their most common IMCI diagnoses was “very serious illness or severe bacterial infection” in 78.8% of cases (Table 2).

Among the 1,786 children aged between 2 to 59 months, 17.5% had received pharmaceutical medication before the IMCI consultation. Main reasons for consultation reported by the accompanying person were: fever (91.4%), cough or breathing difficulties (50.1%); vomiting (35.6%) and diarrhoea (20.5%); 57.7% had fever actually measured by the clinician; 39.8% had a tachycardia, and 55.1% had respiratory signs. Several diagnoses could co-exist for the same child. Their most common IMCI diagnoses were “very severe febrile illness or severe malaria” in 78.4%, followed by “severe pneumonia or very serious illness” in 38.2% of cases.

### Pathways of care and care management according to the severity of hypoxemia

After the index IMCI consultation, the actual referral of severe cases to the district hospital significantly differed according to countries (p<0.001): in Burkina Faso, 73.1% were managed at the PHC, and 26.9% were transferred at hospital. These respective proportions were 88.9% and 10.6% in Guinea, 84.8% and 14.2% in Mali, and 25.3% and 67.0% in Niger. Thus, except in Niger, most of severe cases were managed at PHC level (75.0%), and a small proportion (1.8%) refused care and returned home without receiving any treatment (supplementary figure 2).

Overall, all the following indicators of care management, proportions of referral decision made by clinician, actual hospitalisation, and oxygen therapy for the 142 severe hypoxemia diagnosed (83.8%, 82.3%, and 34.5% respectively) were significantly higher than for the 211 severe cases with moderate hypoxemia (32.7%, 26.5%, and 7.1% respectively), and for the 1,624 severe cases without hypoxemia (26.3%, 17.3%, and 1.4% respectively), indicating an gradient improved case management in case of severe hypoxemia cases compared to moderate hypoxemia, and no hypoxemia (Table 3). The same trend was observed when subgroup analyses were performed for neonates and for children aged between 2 to 59 months.

**Table 3:** Care management of severe cases identified at PHC level in AIRE research sites, and Day-14 mortality according to age groups and level of hypoxemia severity measured at PHC, June 2021 – June 2022; (N=1,998)

|  | Referred to hospital |  | Admitted to hospital |  | Oxygen Therapy |  | Mortality at Day-14 |  |
| --- | --- | --- | --- | --- | --- | --- | --- | --- |
|  | # | % | # | % (95%CI) | # | % | # | % (95%CI) |
| **Global N = 1,998 (100%)** |  |  |  |  |  |  |  |  |
| Severe hypoxemia: 142 (7.1%) | 119 | 83.8 (76.7-89.4) | 117 | 82.3 (75.1-88.3) | 49 | 34.5 (26.7-42.9) | 37 | 26.0 (19.1-34.1) |
| Moderate hypoxemia: 211 (10.6%) | 69 | 32.7 (26.4-39.5) | 56 | 26.5 (20.7-33.0) | 15 | 7.1 (4.0-11.5) | 16 | 7.5 (4.4-12.0) |
| No hypoxemia: 1624 (81.3%) | 426 | 26.3 (24.1-28.4) | 281 | 17.3 (15.5-19.2) | 23 | 1.4 (0.9-2.1) | 38 | 2.3 (1.7-3.2) |
| Children without SpO2 measurement: 21 (1.0) | 11 | 52.4 (29.8-74.3) | 9 | 42.9 (21.8-66.0) | 1 | 4.8 (0.1-23.8) | 4 | 19.0 (5.4-41.9) |
| **Neonates N=212 (10.6%)** |  |  |  |  |  |  |  |  |
| Severe hypoxemia: 32 (15.1%) | 27 | 84.4 (67.2-94.7) | 27 | 84.4 (67.2-94.7) | 15 | 46.9 (29.1-65.3) | 15 | 46.8 (29.1-65.3) |
| Moderate hypoxemia: 42 (19.8%) | 18 | 42.9 (27.7-59.0) | 16 | 38.1 (23.6-54.4) | 5 | 11.9 (4.0-25.6) | 6 | 14.3 (5.4-28.5) |
| No hypoxemia: 131 (61.8%) | 50 | 38.2 (29.8-47.1) | 30 | 22.9 (16.0-31.1) | 6 | 4.6 (1.7-9.7) | 8 | 6.1 (2.7-11.7) |
| Neonates without SpO2 measurement: 7 (3.3) | 5 | 71.4 (29-96.3) | 5 | 71.4 (29-96.3) | 0 | 0.0 (0.0-41.0) | 3 | 42.9 (9.9-81.6) |
| **Children [2-59] N= 1,786 (89.4%)** |  |  |  |  |  |  |  |  |
| Severe hypoxemia:110 (6.2%) | 92 | 83.6 (75.4-90.0) | 90 | 81.8(73.3-88.5) | 34 | 30.9 (22.4-40.4) | 22 | 20.0 (13.0-28.7) |
| Moderate hypoxemia: 169 (9.5%) | 51 | 30.2 (23.4-37.7) | 40 | 23.7(17.5-30.8) | 10 | 5.9 (2.9-10.6) | 10 | 5.9 (2.9-10.6) |
| No hypoxemia: 1,493 (83.5%) | 376 | 25.2 (23-27.5) | 251 | 16.8(14.9-18.8) | 17 | 1.1 (0.7-1.8) | 30 | 2.0 (1.4-2.9) |
| Children without SpO2 measurement: 14 (0.8) | 6 | 42.9 (17.7-71.1) | 4 | 28.6(8.4-58.1) | 1 | 7.1 (0.2-33.9) | 1 | 7.1 (0.2-33.9) |

### Day-14 mortality

Of the 1,998 severe cases included, 1,971 (98.6%) were followed up until Day-14, of whom 95 had died (16 in Burkina Faso, 6 in Guinea, 42 in Mali and 31 in Niger), yielding an overall estimated mortality rate of 4.8% (95%CI: 3.9%-5.8%), with significant between-country heterogeneity: 6.6% (95%CI: 3.8%-10.5%) in Burkina Faso, 1.0% (95%CI: 0.4%-2.1%) in Guinea, 5.2% (95%CI: 3.8%-7.0%) in Mali and 9.6% (95%CI: 6.6%-13.3% in Niger). Overall, 34.0% of deaths occurred among neonates under-2 months of age, with distribution varying between countries: 62.5% (10/16) in Burkina Faso, 33.3% (2/6) in Guinea and 31.0% (13/42) in Mali, and 22.6% (7/31) in Niger.

Day-14 mortality rates for severe hypoxemia cases were the highest, reaching 46.8% for neonates and 20.0% for children aged from 2 to 59 months (Table 3). Overall, hypoxemia was strongly associated with the Kaplan-Meier probability of dying before Day-14, with an increasing gradient according to its severity (log-Rank test; p<0.0001): 26% (95% CI: 19%-33%), 8% (95% CI: 4%-11%), and 3% (95% CI: 2%-3%) for severe hypoxemia, moderate hypoxemia, and no hypoxemia, respectively (Figure 1). Death occurred very early for severe hypoxemia cases (median delay: 1 day). The same trends were observed when stratifying by age groups.

**Figure 1:**
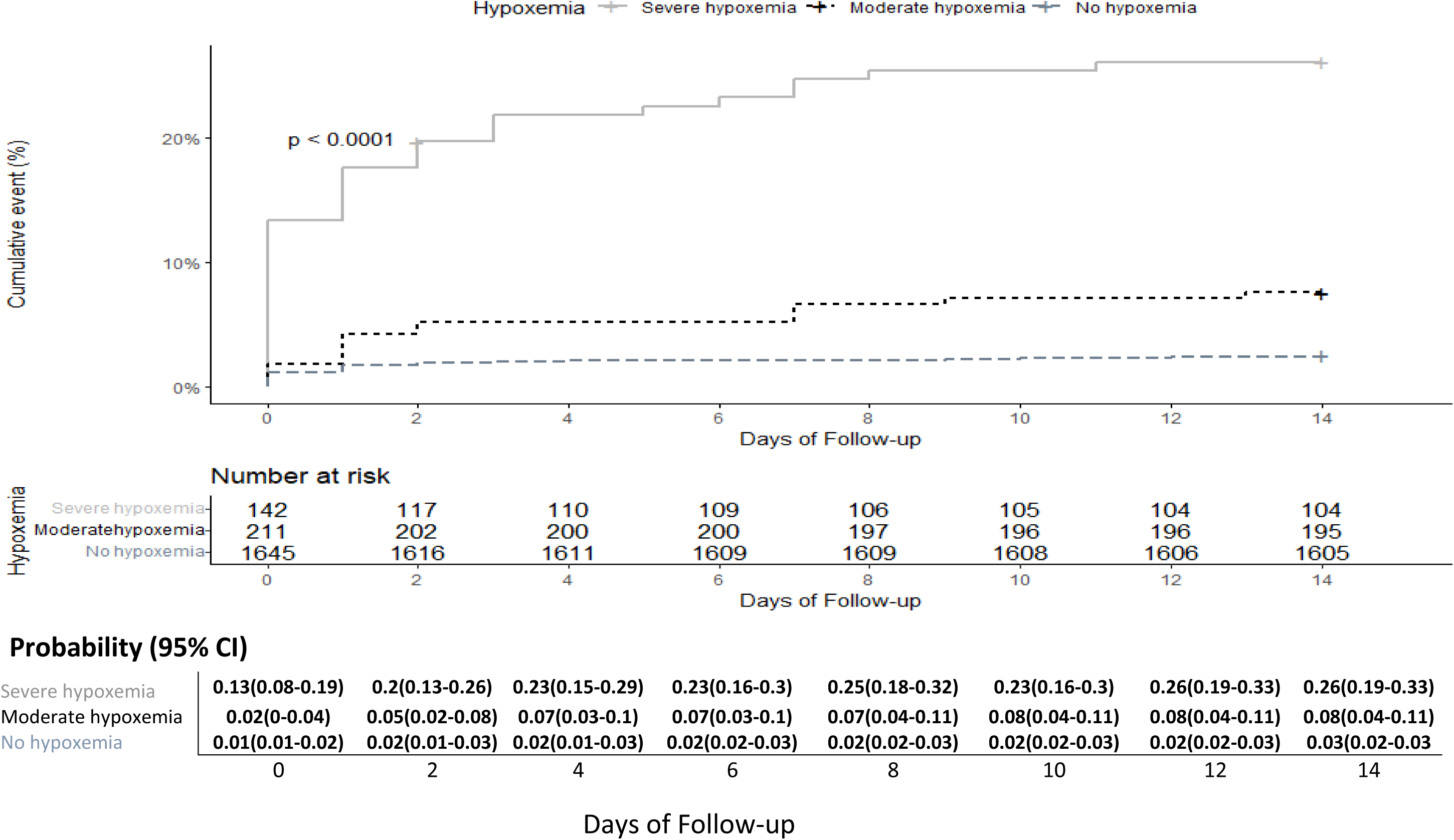
Kaplan Meier curve-Probability of death of severe cases according to the severity of hypoxemia (LogRank test); AIRE research project, 2021 June – 2022 June (N= 1,998)

Overall, the Kaplan-Meier probability of dying before Day-14 varied also significantly with the place of case management (Log-Rank test; p<0.0001): it was significantly higher among severe cases who returned home without care after the IMCI consultation (22%; 95%CI: 7%-35%) and those who were referred to hospital (15%; 95%CI: 11%-18%) compared to those treated at PHC (1.3%; 95%CI: 0.8%-2.0%), (supplementary figure 3). The probability of death reached 17.0% on the same day as the index consultation for those who returned home.

Regardless of the place of initial care management, 55.8% of all deaths occurred at hospital, 18.9% at home, 16.8% during hospital transfer, and 8.4% at the PHC. Deaths occurred early, mainly during hospital transfer or at arrival at the district hospital, with proportions ranging from 64.3% in Mali to 90.3% in Niger, except in Guinea where 83.3% of deaths occurred at PHC (bar plot and Sankey diagram, supplementary figure 4 and 5). Overall, the median delay between the day of IMCI consultation and the day of death for children referred to the district hospital varied from 2.2 days in Niger, to 9.5 days in Guinea. For those who were managed at the PHC level, it ranged from 0.7 days in Guinea to 10.7 days in Burkina Faso.

Regarding the causes of death using the ICD-10 code, malaria was the main diagnosis found in 52.6% of children who died at the PHC level. For those who died at hospital, malaria (33.8%) was also the main diagnosis, followed by acute respiratory infections (16.2%). In Burkina Faso and in Guinea, 31.2% and 33.3% of deaths respectively were respiratory cases while it was estimated at 57.1% in Mali and 61.2% in Niger.

### Predictors of mortality at Day-14

Using a mixed effect Cox regression model to identify predictors of Day-14 mortality for severe cases identified at the PHC level, the following factors were independently associated with an increased risk mortality (Table 4): young age under-2 months (aHR=3.08, 95%CI: 1.41-6.74), moderate hypoxemia (aHR=2.36, 95%CI: 1.23-4.54), severe hypoxemia (aHR=4.23, 95%CI: 2.37-7.54), severe malaria (aHR=2.78, 95%CI: 1.57-4.92), management at hospital (aHR=8.07, 95%CI: 4.46-14.58), and return to home without treatment (aHR=21.33, 95%CI: 8.81-51.61).

**Table 4:** Prognosis factors of Day-14 mortality among the IMCI severe cases identified at PHC level (mixed-effect Cox regression model with a random effect on PHC), (N=95/1,998); AIRE research project, Burkina Faso, Guinea, Mali and Niger - June 2021– June 2022.

| Variables | Levels | Alive | Died | Univariate | Full Adjusted | Final Adjusted* |
| --- | --- | --- | --- | --- | --- | --- |
|  |  |  |  | Fixed effects |  |  |
|  |  |  |  | HR (95% CI, p value) | aHR (95% CI, p value) | aHR (95% CI, p value) |
| Age (months) | [24 - 59] | 915 (48.1) | 25 (26.3) | 1 | 1 | 1 |
|  | <2 | 180 (9.5) | 32 (33.7) | 5.15(3-8.85, p<0.001) | 3.08(1.41-6.74) | 4.5(2.27-8.94) |
|  | [2 - 23] | 808 (42.5) | 38 (40.0) | 1.68(1-2.82, p=0.048)) | 1.27(0.7-2.29) | 1.21(0.7-2.07) |
| Sex | Female | 915 (48.1) | 45 (47.4) | 1 | 1 | 1 |
|  | Male | 988 (51.9) | 50 (52.6) | 1(0.67-1.5, p=0.99) | 1.06(0.68-1.65) | 1.01(0.67-1.53) |
| Ability to read or write of the accompanying | Yes | 459 (24.1) | 20 (21.1) | 1 | Not included |  |
|  | No | 1444 (75.9) | 75 (78.9) | 0.92(0.55-1.55, p=0.76) | Not included |  |
| Income-generating activity of the accompanying | Yes | 545 (28.6) | 17 (17.9) | 1 | 1 |  |
|  | No | 1358 (71.4) | 78 (82.1) | 1.47(0.84-2.58, p=0.17) | 1.03(0.6-1.78) |  |
| Consultation delay since the onset of symptoms (days) | ≤2 | 869 (49.4) | 34 (46.6) | 1 |  |  |
|  | >2 | 891 (50.6) | 39 (53.4) | 1.28(0.8-2.05, p=0.31) | Not included |  |
| Travel delay Home - PHC | ≤30 min | 1387 (72.9) | 61 (64.2) | 1 | 1 |  |
|  | >30 min | 516 (27.1) | 34 (35.8) | 1.71(1.04-2.82, p=0.034) | 0.99(0.62-1.57) |  |
| IMCI Support | Papier-based | 1295 (68.1) | 54 (56.8) | 1 |  |  |
|  | Electronic-based | 608 (31.9) | 41 (43.2) | 1.52(0.62-3.76, p=0.36) | Not included |  |
| Respiratory case | No | 853 (45.4) | 44 (46.8) | 1 | Not included |  |
|  | Yes | 1025 (54.6) | 50 (53.2) | 0.98(0.64-1.5, p=0.93) | - |  |
| Hypoxemia level | No hypoxemia SpO<sub>2</sub> >94 % | 1586 (84.1) | 38 (41.8) | 1 | 1 | 1 |
|  | Moderate SpO<sub>2</sub> [90,93%] | 195(10.3) | 16 (17.6) | 3.32(1.84-5.98, p<0.001) | 2.36(1.23-4.54) | 2.38(1.31-4.33) |
|  | Severe SPO<sub>2</sub> <90% | 105 (5.6) | 37 (40.7) | 10.98(6.83-17.65, p<0.001) | 4.23(2.37-7.54) | 4.26(2.57-7.05) |
| Heart rate according to age | Normal | 1387 (78.5) | 62 (75.6) | 1 | 1 |  |
|  | Bradycardia | 52 (2.9) | 12 (14.6) | 5.72(3.03-10.81, p=0) | 1.83(0.93-3.57, p=0.079) |  |
|  | Tachycardia | 329 (18.6) | 8 (9.8) | 0.87(0.53-1.43, p=0.58) | 0.97(0.58-1.61, p=0.89) |  |
| Severe pneumonia | No | 1236 (65.0) | 80 (84.2) | 1 | 1 |  |
|  | Yes | 667 (35.0) | 15 (15.8) | 0.56(0.3-1.04, p=0.067) | 0.68(0.37-1.26) |  |
| Severe malaria | No | 1119 (58.8) | 58 (61.1) | 1 | 1 |  |
|  | Yes | 784 (41.2) | 37 (38.9) | 0.6(0.37-0.97, p=0.036) | 2.78(1.57-4.92) | 3.06(1.79-5.25) |
| Severe acute malnutrition | No | 1726 (90.7) | 81 (85.3) | 1 | Not included |  |
|  | Yes | 177 (9.3) | 14 (14.7) | 1.15(0.63-2.1, p=0.66) |  |  |
| Place of care management | PHC | 1480 (77.8) | 19 (20.0) | 1 | 1 | 1 |
|  | Hospital | 395 (20.8) | 68 (71.6) | 13.89(8.15-23.67, p<0.001) | 8.07(4.46-14.58) | 8.16(4.6-14.47) |
|  | Back to home | 28 (1.5) | 8 (8.4) | 24.71(10.47-58.35, p<0.001) | 21.33(8.81-51.61) | 16.54(7-39.08) |
| aHR: adjusted Hazard Ratio; *Multivariate model estimated using a top-down step-by-step procedure. |  |  |  |  |  |  |

## Discussion

To our knowledge, this is the first cohort providing short-term follow-up data on the prevalence, care management, and mortality outcome of critically ill under-5 children according to hypoxemia level identified in primary care using IMCI and PO in four West African countries. Our results highlighted some specifics findings as follows.

Firstly, overall hypoxemia was frequent among critically ill children at primary care reaching, 17.6% (7.1% severe and 10.5% moderate). Severe hypoxemia affected both respiratory (9.9%) and non-respiratory (3.7%) cases. High severe hypoxemia was similarly reported among severe cases in Papua New Guinea but higher at hospital admission reaching 73% and 32% of those with and without acute low respiratory illness, respectively (27). Severe hypoxemia was also significantly higher among youngest children compared to older ones: this is consistent with the findings reported among hospitalized children in Nigeria: 22.2% for neonates vs 10.2% for children (8) and in a study conducted in Malawi through PO use among 27,586 children under-5 with clinical pneumonia seeking seven hospitals, 18 health centres, and with 38 community health workers (11.4% for 0–5 months old, 8.4% for 6–23 month olds and 4.7% for 24–59 month olds) (28).

Secondly, our study highlighted that the use of PO integrated to IMCI guidelines at the PHC level allowing for the diagnosis of hypoxemia has improved the case management of severe cases, as referral decision, effective hospitalization, and oxygen therapy rates were all significantly higher for severe hypoxemic children than for those with moderate or no hypoxemia. Despite our uncontrolled design, this gradient observed in the relationship between the severity of hypoxemia and the improved of care management arguing for causal effect of PO use, that will be detailed elsewhere. Similarly, McCollum et al. already reported in 2016 that Malawian outpatient children with severe hypoxemia were significantly more than twice as likely to have been referred as those with SpO2, 84.3% (385/457) vs 41.5% (871/2099) (21). The use of pulse oximetry has also increased the referral rate of severely hypoxemic but asymptomatic children, without chest indrawing and without signs of danger, from 0% to 27.2%. In an Ethiopian cluster-randomized trial, the diagnosis of hypoxemia using PO also prompted the referral rate in severe cases (23). We assume that PO has improved the clinicians’ decision in case management through self-confidence, and assurance in the diagnosis made as reported in our study (29) and elsewhere (30,31). It was also observed by Rahman et al. in Bangladesh in 2022, where HCW and parents were in favour of using PO to assess children (32).

Thirdly, despite the WHO IMCI recommendations to refer urgently severe cases, we observed an overall very low rate of hospital referral (<25%) of those identified at PHC using IMCI+PO, except in Niamey (Niger), an urban area where population was very close to the district hospital. More specifically, many severe cases with severe hypoxemia remained managed at PHC level, where no oxygen is available. As we selected research PHC in the most favourable situation regarding the capacity of referral (existing ambulance, community support) (24,33), we supposed that although low, we may have over-estimated this referral capacity at the country level. This low compliance with hospital referral concur with those consistently reported in other contexts: In Ethiopia, only 37% of young infants and 50% of children aged from 2 to 59 months at primary care who deserved an urgent hospital referral were referred (34). In Papua New Guinea, among the 139 outpatients severe cases identified, only 60% resulted in hospitalization in 2019 (35). In Uganda, only 44% (12/27) of severe hypoxemic cases diagnosed at PHC were referred (7). In Malawi that even though the hospital referral rate was better among children for whom PO had been used, all the severe cases had not been referred

(21). Several contextual factors could explain these low referral rates. The main one is related to poverty with most of families unable to pay the referral fees, such as in West Africa. Indeed, we reported elsewhere that referral to hospital is associated with higher expenses than at the PHC (36), and also with the cessation of income-generating activities, with a loss of resources and disorganization of the family unit, as there are often other children left at home. Some parents may also refuse hospital transfer. This was also reported by Graham *et al*. in Uganda, where 16% (8/51) of families were unable to pay (7). Faced with the child’s poor clinical manifestation, parents may be unconvinced of the severity of the disease and refuse the clinician’s decision of referral, thinking it’s unnecessary (7,37), or on the basis of their previous experience of oxygen therapy (38). More specifically in the AIRE project, we faced also the West African specificities of the national IMCI management programs for some specific severe diseases, such as malaria or malnutrition without complications, allowing those to be treated at the PHC level (unpublished national health protocols). In addition, marabouts, fetishists, as well as traditional healers can also hinder access to care. A study analysing causal pathways for children dying from treatable illnesses in Mozambique shown that the seeking delay was significantly higher among the children who were taken to a traditional healer, (39). In a study, conducted in rural Mali, traditional healers are a barriers in seeking care for 8% of children under-5 (40). Finally, clinicians sometimes did not refer severe cases due to non-functional ambulances (41), or because of the road insecurity, or geographical barriers (mountains, hills, rivers and long distances) (7,39,42–44).

Fourthly, the access to oxygen therapy remained insufficient. In our study, only 34.5% of severe cases with hypoxemia identified at PHC level actually received oxygen, and their mortality exceed 26.0% signing a late referral. In Malawi, 22.5% of severely hypoxemic children identified at PHC received oxygen (45) ; it was 12% in Uganda (7). This low rate of oxygen therapy among children who need it reveals the difficulties of access to oxygen, which may be due to a lack of supply (46) to the unavailability of oxygen or even breaks in supply at hospital level, but also to the lack of affordability for patients. In 2021, in Nigeria, only 59% of the health facilities had a functional source of oxygen available on the day of inspection, and oxygen services were expensive in hospitals and private facilities (47). Finally, this lack of access can be related to a lack of knowledge on oxygen use on the part of HCW, who were not or were poorly trained (48), and to the lack of ambulances or adequate and efficient hospital referral systems. Our results highlighted the major issue of access to oxygen in most African countries, particularly revealed by the Covid-19 pandemic (49,50). Several initiatives are being set up in conjunction with Africa Centres for Diseases Control and Prevention, and other stakeholders to make oxygen available on a sustainable basis (51,52), including at the peripheral level. Graham et al shown that large-scale improvements in hospital oxygen services could have the potential to improve clinical outcomes (53).

Fifthly, in our study, Day-14 mortality risk was high, and correlated with the severity of hypoxemia for all children with SpO2≤93%. This mortality occurred early, and the cases ultimately referred to hospital were very serious ones, signing a late and lack referral to hospital. Day-14 mortality was also significantly higher for those referred and managed in district hospital, and those who returned home without care compared to those managed at PHC level. Several barriers refraining access to care could explained the high Day-14 mortality observed: delayed consultation (42,54–56), delayed hospital transfer as well as the difficult transfer conditions (41) or the absence or inadequate treatment during transfer as many deaths occur during transfer to hospital. According to our data from the baseline assessment of PHCs in the AIRE project run in 2020 (33), only 16.3% of all PHCs assessed had a functional ambulance of which only two had oxygen on board. Both severe and moderate hypoxemia were independent predictor of death among severely ill children. According to Graham in 2019, hypoxemic newborns and hypoxemic children had a 6-fold and 7-fold greater risk of death respectively compared to non-hypoxemic children (8). Hooli in 2023 in a systematic review demonstrated that severe hypoxemia presented 3.4 times greater risk of death in hospitalized children with pneumonia with chest indrawing (57). It was also consistent with a review of risk factors for death in pneumonia cases reported in 2023 (58). Therefore, our results strongly support the question of using a higher SpO2 level at PHC level to recommend hospital referral to provide earlier access to oxygen for hypoxemic children, as reported by Graham *et al* in 2024 (59). This major point deserves to be evaluated in further research and could be taken into consideration when updating the WHO guidelines recommending a higher threshold for hospital referral of hypoxemic children different from the threshold for oxygen administration.

Overall, our study presents several limitations: first, the definition of severe cases based on IMCI guidelines, is slightly different in each country, especially in Guinea for cases of pneumonia. This could lead to a bias in overestimation of severe cases in Guinea, that needs to be considered when comparing data between countries. Another limitation lies in the absence of etiological diagnosis other than mRDT leading to imprecise or inaccurate clinical diagnosis, relying on the clinician’s expertise and knowledge. This gap was partially addressed during the initial training supervisions jointly organized with the project and Ministries of Health staffs, aimed at improving IMCI practice using PO and current management protocols. We carried out this non randomised operational research, under field conditions, to assess the PO introduction very pragmatically, according to the clinician’s decision of case management. In addition, the selection of AIRE intervention health districts and research PHCs (24) made them unrepresentative of all PHCs at national level. This would lead to a bias in the selection of severe cases and subsequent difficulties in statistical inference, but the results of this study have the advantage of informing health authorities and guiding them, based on scientific evidence, on the best decision to take for the health of populations. Our study has also strengths: the data collected as part of this large sample size, including neonates are valuable and rare, especially care pathway data. Very few studies have examined the same topic in the West African context. The quality of the data collected, which is almost complete, with very few missing data also constitute a strong point of the AIRE study. We recorded a low rate of lost to follow-up of around 1.4%, which is an indicator of good follow-up of the cohort of severe cases, especially since the follow-up was carried out at community level despite difficult field conditions.

## Conclusion

Our study, conducted at primary care level in West-African under-5 children, highlighted that hypoxemia is high among severe cases, but many including hypoxemic severe cases remained managed at PHC level, where oxygen was not available. Nevertheless, the use of PO improved children’s case management, particularly for those with severe hypoxemia, but there were difficulties accessing oxygen at the hospital level. The Day-14 mortality was high especially for children who were managed at district hospital level marking their delayed care management. Both severe and moderate hypoxemia were independent factors of Day-14 mortality in critically ill children under-5, supporting the need to consider higher SpO2 level at primary care to indicate hospital referral. To improve the survival of severe cases identified at PHC level including those with severe hypoxemia, West African governments must pay attention to costs associated with transport and care at the hospital, support earlier and effective hospital transfer and provide access to oxygen.

## Data Availability

All data produced in the present study are available upon reasonable request to the authors

---

**Supplementary figure 1:**
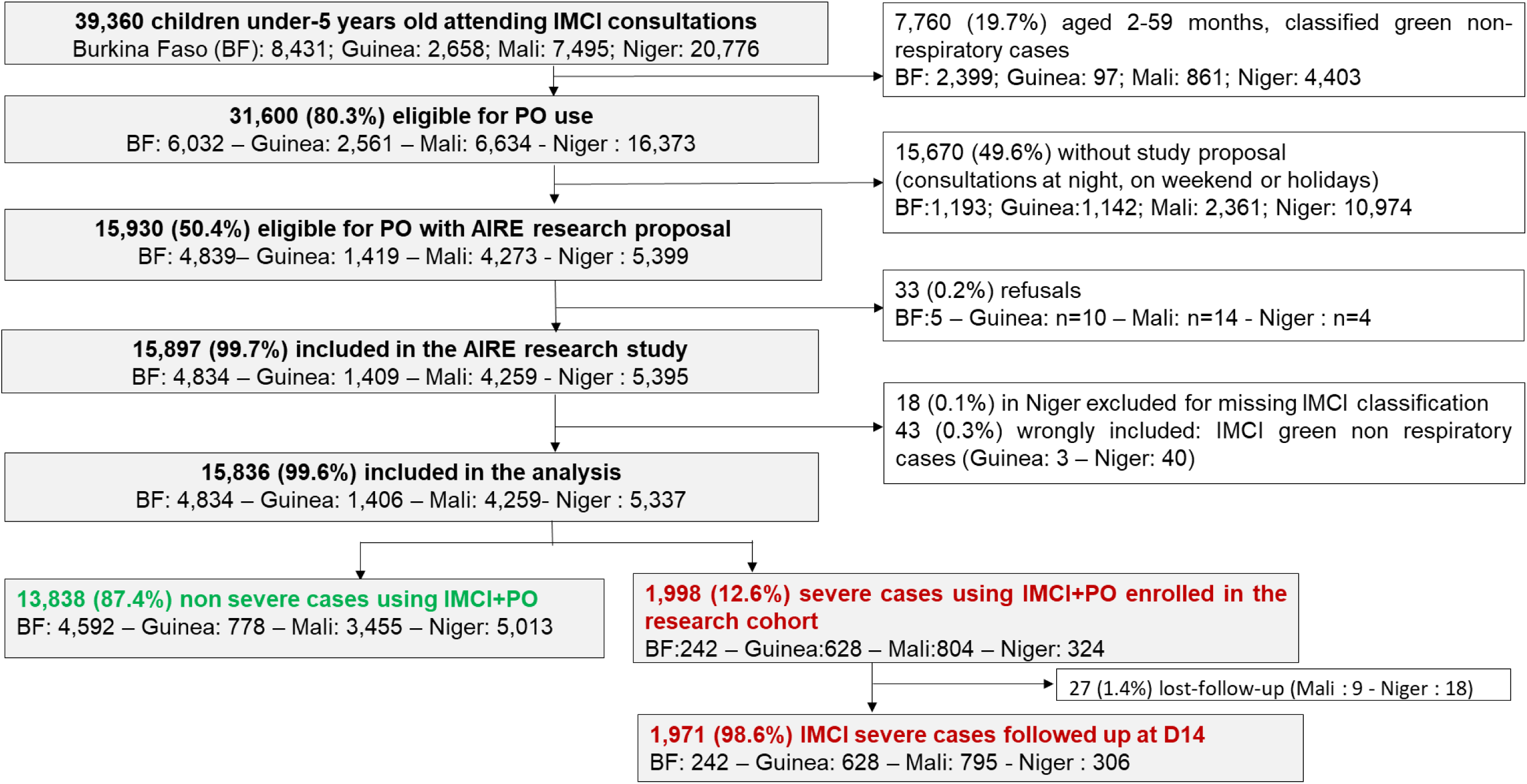
Flowchart of the inclusion process of the AIRE research project. June 2021– June 2022

**Supplementary figure 2:**
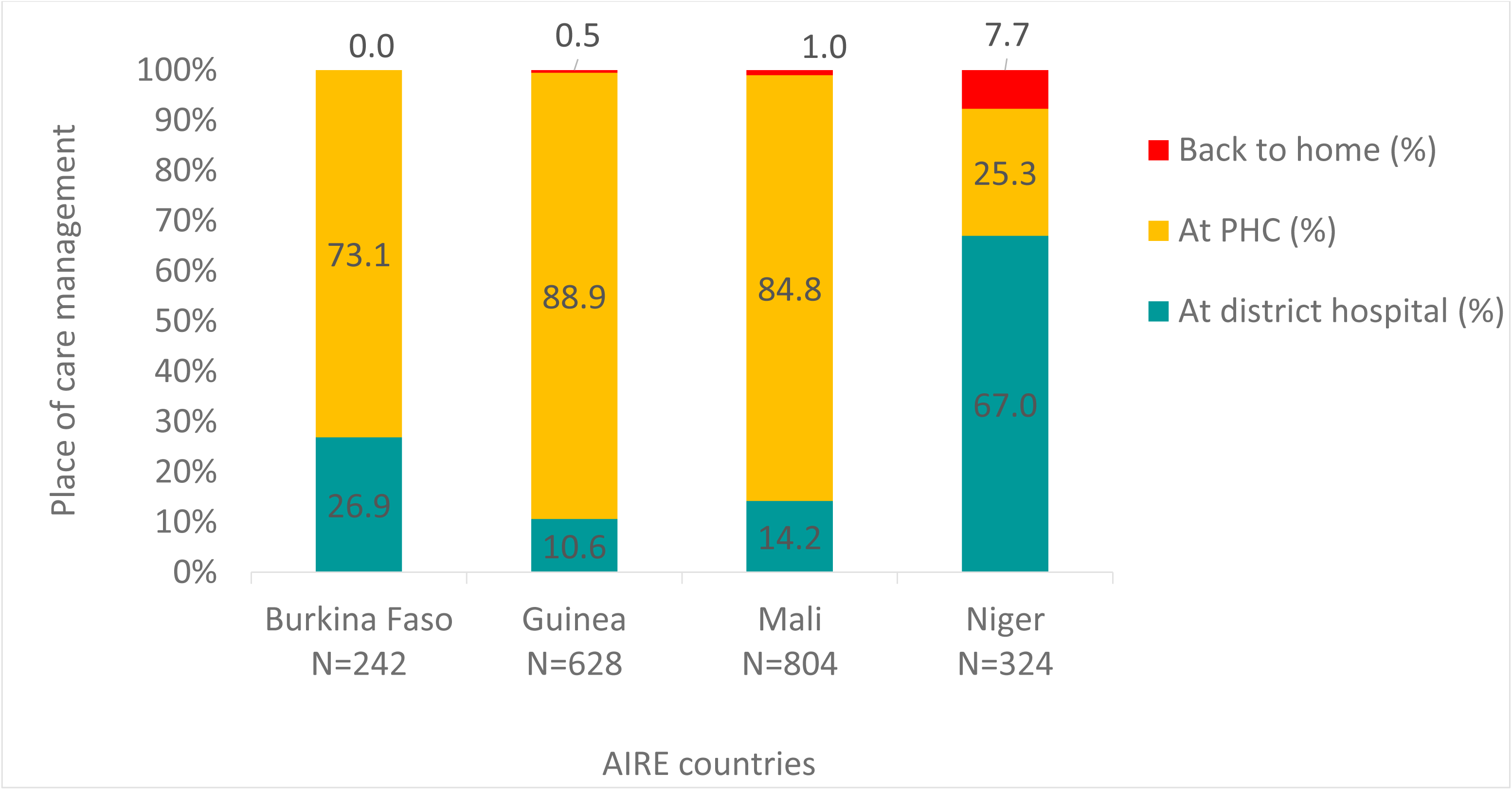
Place of care management of severe cases by country in the AIRE research project, 2021 June – 2022 June (N=1,998)

**Supplementary figure 3:**
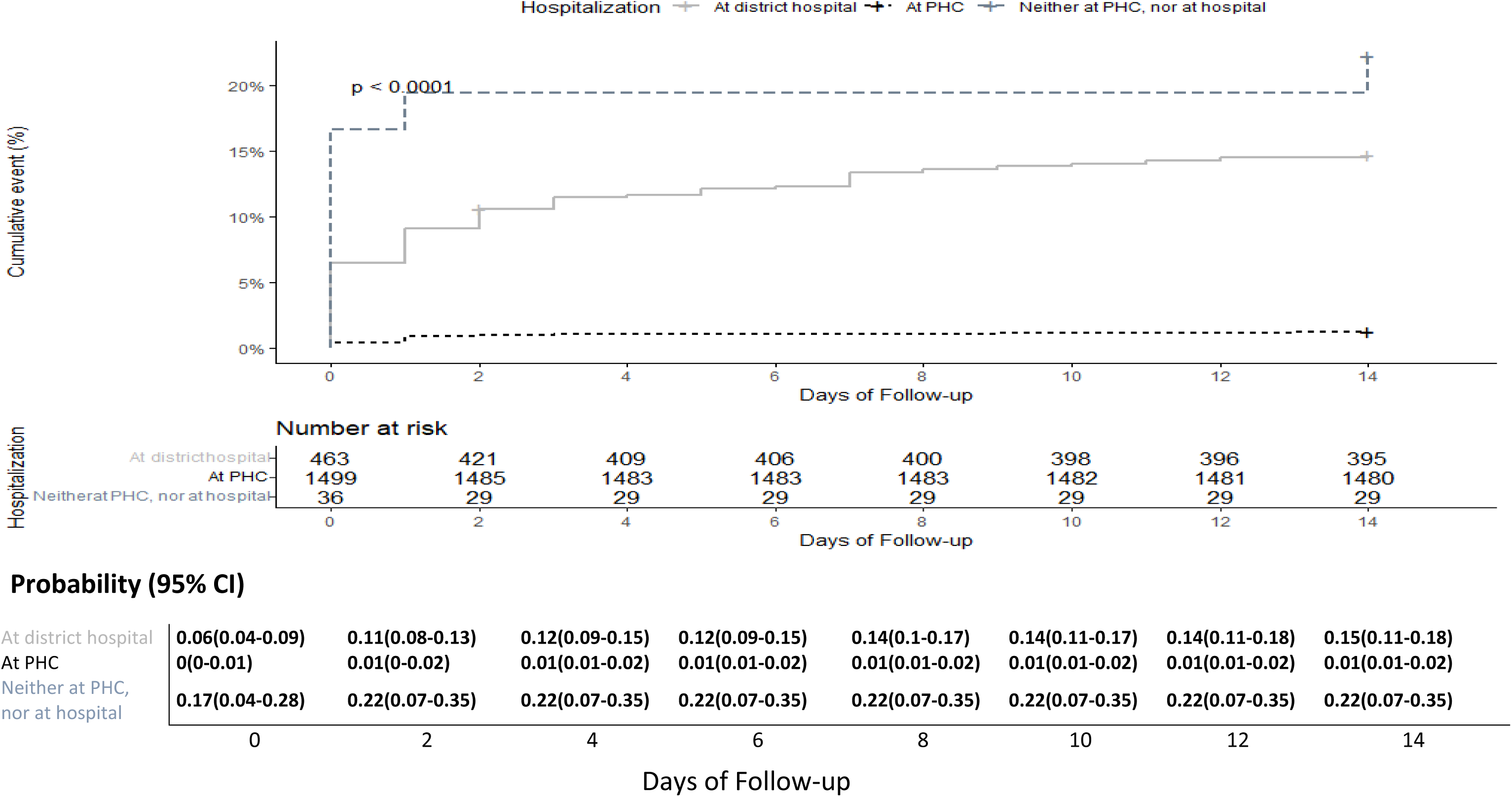
Kaplan Meier probability of death of severe cases according to the place of care management (LogRank test); AIRE research 2021 June – 2022 June (N= 1,998)

**Supplementary Figure 4:**
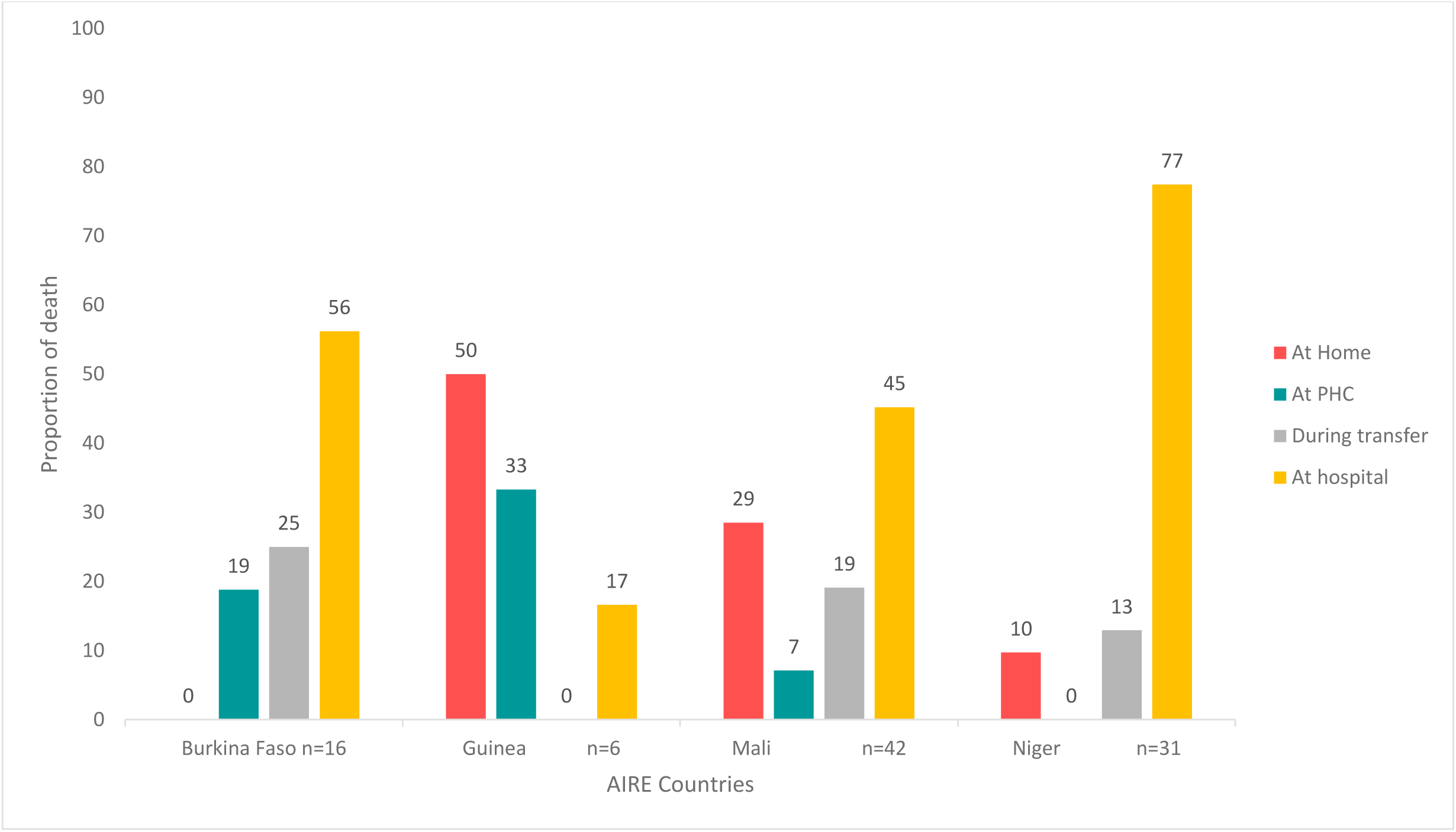
Place of death of IMCI+PO severe cases identified at PHC level in the AIRE research project, June 2021– June 2022 (N=95/1,998)

**Supplementary figure 5:**
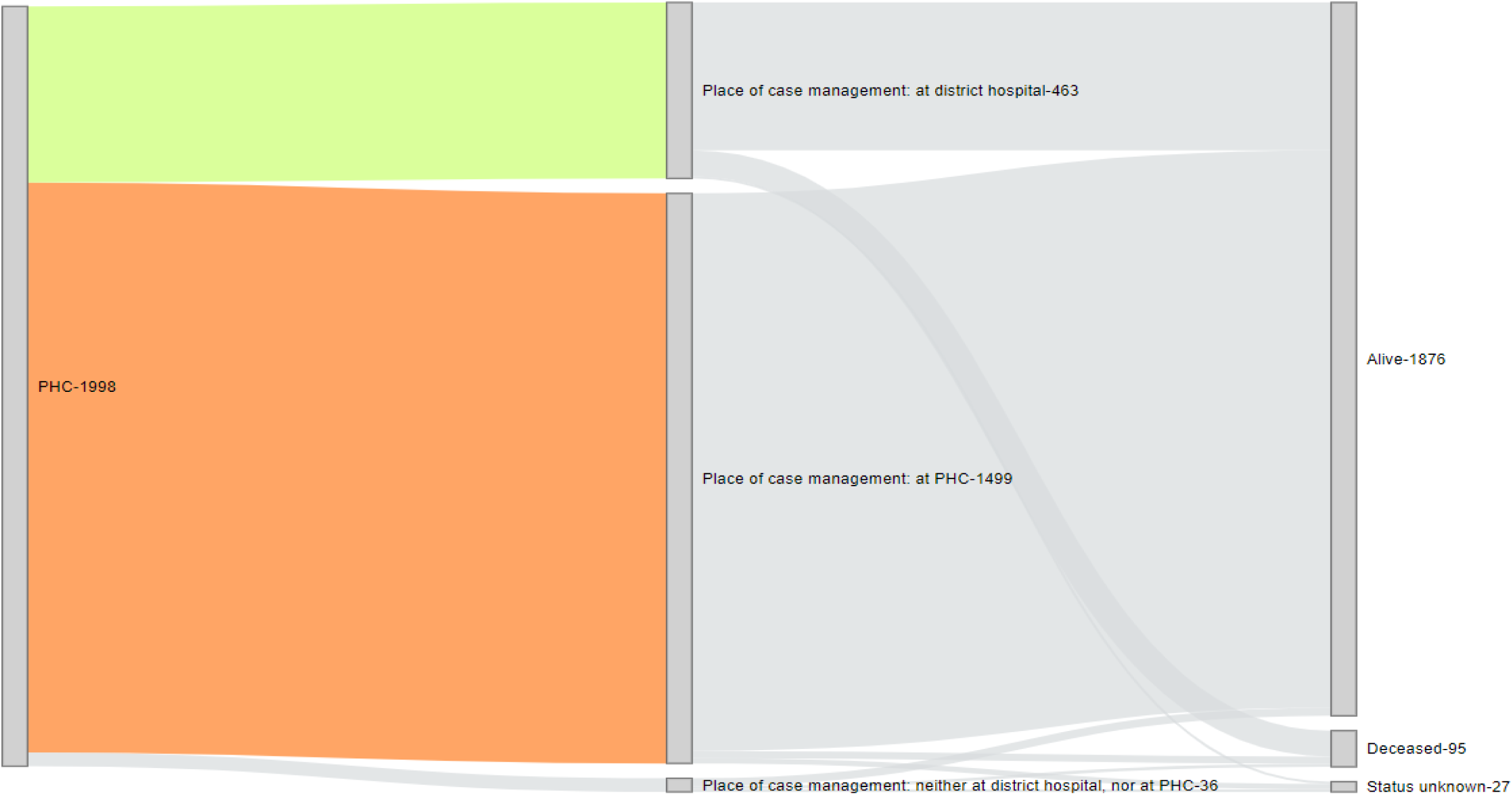
Sankey plot showing the pathway of IMCI+PO severe cases identified at PHC and followed up until Day-14 in the AIRE research study, June 2021- June 2022, (N)

## Appendix

## ACKNOWLEDGMENTS

We thank all the children and their families who participated in the study, as well as the healthcare staff at the participating hospitals and the sites involved. We thank the field project staff and the AIRE Research Study Group*. We thank the Ministries of Health of the participating countries for their support.

**Acknowledgements: Children, families, UNITAID and The AIRE Research Study Group: Country investigators**: Ouagadougou, Burkina Faso: S. Yugbaré Ouédraogo (PI), V. M. Sanon Zombré (CoPI), Conakry, Guinea: M. Sama Cherif (CoPI), I. S. Diallo (CoPI), D. F. Kaba, (PI). Bamako, Mali: A. A. Diakité (PI), A. Sidibé, (CoPI). Niamey, Niger: H. Abarry Souleymane (CoPI), F. Tidjani Issagana **Dikouma (PI).** Research coordinators & data centers: Inserm U1295, Toulouse 3 University, France: H. Agbeci (Int Health Economist), L. Catala (Research associate), D. L. Dahourou (Research associate), S. Desmonde (Research associate), E. Gres (PhD Student), G. B. Hedible (Int research project manager), V. Leroy (research coordinator), L. Peters Bokol (Int clinical research monitor), J. Tavarez (Research project assistant), Z. Zair (Statistician, Data scientist). **CEPED, IRD, Paris, France:** S. Louart (process manager), V. Ridde (process coordination). **Inserm U1137, Paris, France:** A. Cousien (Research associate). **Inserm U1219,** EMR271 IRD, **Bordeaux University, France**: R. Becquet (Research associate), V. Briand (Research associate), V. Journot (Research associate). **PACCI, CHU Treichville, Abidjan, Côte d’Ivoire**: S. Lenaud (Int data manager), C. N’Chot (Research associate), B. Seri (Supervisor IT), C. Yao (data manager supervisor). **Consortium NGOs partners: Alima-HQ (consortium lead), Dakar, Sénégal**: G. Anago (Int Monitoring Evaluation Accountability And Learning Officer), D. Badiane (Supply chain manager), M. Kinda (Director), D. Neboua (Medical officer), P. S. Dia (Supply chain manager), S. Shepherd (referent NGO), N. di Mauro (Operations support officer), G. Noël (Knowledge broker), K. Nyoka (Communication and advocacy officer), W. Taokreo (Finance manager), O. B. Coulidiati Lompo (Finance manager), M. Vignon (Project Manager). **Alima, Conakry, Guinea:** P. Aba (clinical supervisor), N. Diallo (clinical supervisor), M. Ngaradoum (Medical Team Leader), S. Léno (data collector), A. T. Sow (data collector), A. Baldé (data collector), A. Soumah (data collector), B. Baldé (data collector), F. Bah (data collector), K. C. Millimouno (data collector), M. Haba (data collector), M. Bah (data collector), M. Soumah (data collector), M. Guilavogui (data collector), M. N. Sylla (data collector), S. Diallo (data collector), S. F. Dounfangadouno (data collector), T. I. Bah (data collector), S. Sani (data collector), C. Gnongoue (Monitoring Evaluation Accountability And Learning Officer), S. Gaye (Monitoring Evaluation Accountability And Learning Officer), J. P. Y. Guilavogui (Clinical Research Assistant), A. O. Touré (Country health economist), J. S. Kolié (Country clinical research monitor), A. S. Savadogo (country project manager). **Alima, Bamako, Mali:** F. Sangala (Medical Team Leader), M. Traore (Clinical supervisor), T. Konare (Clinical supervisor), A. Coulibaly (Country health economist), A. Keita (data collector), D. Diarra (data collector), H. Traoré (data collector), I. Sangaré (data collector), I. Koné (data collector), M. Traoré (data collector), S. Diarra (data collector), V. Opoue (Monitoring Evaluation Accountability And Learning Officer), F. K. Keita (medical coordinator), M. Dougabka (Clinical research assistant then Monitoring Evaluation Accountability And Learning Officer), B. Dembélé (data collector then Clinical research assistant), M. S. Doumbia (country health economist), G. D. Kargougou (country clinical research monitor), S. Keita (country project manager). **Solthis-HQ, Paris**: S. Bouille (NGO referent), S. Calmettes (NGO referent), F. Lamontagne (NGO referent). **Solthis, Niamey:** K. H. Harouna (clinical supervisor), B. Moutari (clinical supervisor), I. Issaka (clinical supervisor), S. O. Assoumane (clinical supervisor), S. Dioiri (Medical Team Leader), M. Sidi (data collector), K. Sani Alio (Country supply chain officer), S. Amina (data collector), R. Agbokou (Clinical research assistant), M. G. Hamidou (Clinical Research Assistant), S. M. Sani (Country health economist), A. Mahamane, Aboubacar Abdou (data collector), B. Ousmane (data collector), I Kabirou (data collector), I. Mahaman (data collector), I Mamoudou (data collector), M. Baguido (data collector), R. Abdoul (data collector), A. Sahabi (data collector), F. Seini (data collector), Z. Hamani (data collector), L-Y B Meda (Country clinical research monitor), Mactar Niome (country project manager), X. Toviho (Monitoring Evaluation Accountability And Learning Officer), I. Sanouna (Monitoring Evaluation Accountability And Learning Officer), P. Kouam (program officer). **Terre des hommes-HQ, Lausanne:** S. Busière (NGO referent), F. Triclin (NGO referent). **Terre des hommes, BF:** A. Hema (country project manager), M. Bayala (IeDA IT), L. Tapsoba (Monitoring Evaluation Accountability And Learning Officer), J. B. Yaro (Clinical reearch assistant), S. Sougue (Clinical reearch assistant), R. Bakyono (Country health economist), A. G. Sawadogo (Country clinical research monitor), A. Soumah (data collector), Y. A. Lompo (data collector), B. Malgoubri (data collector), F. Douamba (data collector), G. Sore (data collector), L. Wangraoua (data collector), S. Yamponi (data collector), S. I. Bayala (data collector), S. Tiegna (data collector), S. Kam (data collector), S. Yoda (data collector), M. Karantao (data collector), D. F. Barry (Clinical supervisor), O. Sanou (clinical supervisor), N. Nacoulma (Medical Team Leader), N. Semde (clinical supervisor), I. Ouattara (Clinical supervisor), F. Wango (clinical supervisor), Z. Gneissien (clinical supervisor), H. Congo (clinical supervisor). **Terre des hommes, Mali:** Y. Diarra (clinical supervisor), B. Ouattara (clinical supervisor), A. Maiga (data collector), F. Diabate (data collector), O. Goita (data collector), S. Gana (data collector), S. Diallo (data collector), S. Sylla (data collector), D. Coulibaly (Tdh project manager), N. Sakho (NGO referent). **Country SHS team: Burkina Faso**: K. Kadio (consultant and research associate), J. Yougbaré (data collector), D. Zongo (data collector), S. Tougouma (data collector), A. Dicko (data collector), Z. Nanema (data collector), I. Balima (data collector), A. Ouedraogo (data collector), A. Ouattara (data collector), S. E. Coulibaly (data collector). **Guinea**: H. Baldé (consultant and research associate), L. Barry (data collector), E. Duparc Haba (data collector). **Mali**: A. Coulibaly (consultant and research associate), T. Sidibe (data collector), Y. Sangare (data collector), B. Traore (data collector), Y. Diarra (data collector). **Niger**: A. E. Dagobi (consultant and research associate), S. Salifou (data collector), B. Gana Moustapha Chétima (data collector), I. H. Abdou (data collector)

## FOOTNOTES

### Handling editor

#### Contributors

VL and VR conceptualised the research. The AIRE Research Study Group conducted training, data collection and management. ZZ with contributions from HGB and VL realise the data analysis. HGB prepared the first draft of this article. All authors were involved in data interpretation and review of the final manuscript. VL is the guarantor to submit the manuscript.

#### Funding

The AIRE project is funded by UNITAID, with in-kind support from Inserm and IRD. UNITAID was not involved in the design of the study, the collection, analysis and interpretation of the data, nor in the writing of the manuscript.

#### Ethics approval and consent to participate

Ethics approval and consent to participate The AIRE research protocol, the information notice (translated in vernacular languages), the written consent form and any other relevant document have been submitted to each national ethics committee, to the Inserm Institutional Evaluation Ethics Committee (IEEC) and to the WHO Ethics Review Committee (WHO–ERC). All the aforementioned ethical committees reviewed and approved the protocol and other key documents (Comité d’Ethique pour la Recherche en Santé (CERS), Burkina Faso n°2020–4–070; Comité National d’Ethique pour la Recherche en Santé (CNERS), Guinea n°169/CNERS/21; Comité National d’Éthique pour la Santé et les Sciences de la vie (CNESS), Mali n°127/MSDS–CNESS; Comité National d’Ethique pour la Recherche en Santé (CNERS) Niger n°67/2020/CNERS; Inserm IEEC n°20–720; WHO–ERC n° ERC.0003364). This study has been retrospectively registered by the Pan African Clinical Trials Registry on June 15th 2022 under the following Trial registration number: PACTR202206525204526.

#### Data Availability Statement

The datasets generated and analysed during the current study are not publicly available. Access to processed deidentified participant data will be made available to any third Party after the publication of the main AIRE results stated in the Pan African Clinical Trial Registry Study statement (PACTR202206525204526, registered on 06/15/2022), upon a motivated request (concept sheet), and after the written consent of the AIRE research coordinator (Valeriane Leroy, Inserm U1295 Toulouse, France, orcid.org/0000-0003-3542-8616) obtained after the approval of the AIRE publication committee, if still active.

## References

1. World Health Organization. Enfants : améliorer leur survie et leur bien-être [Internet]. 2020 [cité 8 sept 2023]. Disponible sur: https://www.who.int/fr/news-room/fact-sheets/detail/children-reducing-mortality

2. UN-IGME-Child-Mortality-Report-2022.pdf [Internet]. [cité 23 août 2023]. Disponible sur: https://childmortality.org/wp-content/uploads/2023/01/UN-IGME-Child-Mortality-Report-2022.pdf

3. Unicef. Scribd. 2016 [cité 10 sept 2023]. UNICEF Pneumonia Diarrhoea Report2016 Web Version - Final PDF | PDF | Diarrhea | Pneumonia. Disponible sur: https://www.scribd.com/document/403944138/UNICEF-Pneumonia-Diarrhoea-report2016-web-version-final-pdf

4. Unicef. Levels and Trends in Child Mortality Report 2018 | UNICEF [Internet]. 2018 [cité 10 sept 2023]. Disponible sur: https://www.unicef.org/reports/levels-and-trends-child-mortality-report-2018

5. Wang H, Abajobir AA, Abate KH, Abbafati C, Abbas KM, Abd-Allah F, et al. Global, regional, and national under-5 mortality, adult mortality, age-specific mortality, and life expectancy, 1970– 2016: a systematic analysis for the Global Burden of Disease Study 2016. The Lancet. 16 sept 2017;390(10100):1084–150.

6. World Health Organization. Oxygen therapy for children: a manual for health workers [Internet]. Geneva: World Health Organization; 2016 [cité 24 août 2023]. 57 p. Disponible sur: https://apps.who.int/iris/handle/10665/204584

7. Graham HR, Kamuntu Y, Miller J, Barrett A, Kunihira B, Engol S, et al. Hypoxaemia prevalence and management among children and adults presenting to primary care facilities in Uganda: A prospective cohort study. PLOS Glob Public Health. 22 avr 2022;2(2):e0000352.

8. Graham H, Bakare AA, Ayede AI, Oyewole OB, Gray A, Peel D, et al. Hypoxaemia in hospitalised children and neonates: A prospective cohort study in Nigerian secondary-level hospitals. eClinicalMedicine. 1 nov 2019;16:51–63.

9. Hooli S, King C, Zadutsa B, Nambiar B, Makwenda C, Masache G, et al. The Epidemiology of Hypoxemic Pneumonia among Young Infants in Malawi. Am J Trop Med Hyg. mars 2020;102(3):676–83.

10. Ashley EA, Poespoprodjo JR. Treatment and prevention of malaria in children. Lancet Child Adolesc Health. 1 oct 2020;4(10):775–89.

11. Bhatt S, Weiss DJ, Cameron E, Bisanzio D, Mappin B, Dalrymple U, et al. The effect of malaria control on Plasmodium falciparum in Africa between 2000 and 2015. Nature. 8 oct 2015;526(7572):207–11.

12. Murray GPD, Lissenden N, Jones J, Voloshin V, Toé KH, Sherrard-Smith E, et al. Barrier bednets target malaria vectors and expand the range of usable insecticides. Nat Microbiol. janv 2020;5(1):40–7.

13. Bergeron G, Castleman T. Program Responses to Acute and Chronic Malnutrition: Divergences and Convergences123. Adv Nutr. 2 mars 2012;3(3):242–9.

14. Ireen S, Raihan MJ, Choudhury N, Islam MM, Hossain MI, Islam Z, et al. Challenges and opportunities of integration of community based Management of Acute Malnutrition into the government health system in Bangladesh: a qualitative study. BMC Health Serv Res. 10 avr 2018;18:256.

15. World Health Organization et UNICEF. Handbook : IMCI integrated management of childhood illness [Internet]. World Health Organization et UNICEF; 2005 [cité 13 sept 2023]. Disponible sur: https://apps.who.int/iris/handle/10665/42939

16. World Health Organization. Integrated Management of Childhood Illness: distance learning course [Internet]. Geneva: World Health Organization; 2014 [cité 25 oct 2023]. 15 p. Disponible sur: https://iris.who.int/handle/10665/104772

17. World Health Organization. Revised WHO classification and treatment of pneumonia in children at health facilities: evidence summaries [Internet]. Geneva: World Health Organization; 2014 [cité 10 sept 2023]. 26 p. Disponible sur: https://apps.who.int/iris/handle/10665/137319

18. Cunningham J, Jones S, Gatton ML, Barnwell JW, Cheng Q, Chiodini PL, et al. A review of the WHO malaria rapid diagnostic test product testing programme (2008–2018): performance, procurement and policy. Malar J. 2 déc 2019;18:387.

19. Millar KR, McCutcheon J, Coakley EH, Brieger W, Ibrahim MA, Mohammed Z, et al. Patterns and predictors of malaria care-seeking, diagnostic testing, and artemisinin-based combination therapy for children under five with fever in Northern Nigeria: a cross-sectional study. Malar J. 21 nov 2014;13:447.

20. Lim YW, Steinhoff M, Girosi F, Holtzman D, Campbell H, Boer R, et al. Reducing the global burden of acute lower respiratory infections in children: the contribution of new diagnostics. Nature. nov 2006;444(1):9–18.

21. McCollum ED, King C, Deula R, Zadutsa B, Mankhambo L, Nambiar B, et al. Pulse oximetry for children with pneumonia treated as outpatients in rural Malawi. Bull World Health Organ. 1 déc 2016;94(94):893–902.

22. Boyd N, King C, Walker IA, Zadutsa B, Bernstein M, Ahmed S, et al. Usability Testing of a Reusable Pulse Oximeter Probe Developed for Health-Care Workers Caring for Children < 5 Years Old in Low-Resource Settings. Am J Trop Med Hyg. oct 2018;99(4):1096–104.

23. Tesfaye SH, Gebeyehu Y, Loha E, Johansson KA, Lindtjørn B. Pulse oximeter with integrated management of childhood illness for diagnosis of severe childhood pneumonia at rural health institutions in Southern Ethiopia: results from a cluster-randomised controlled trial. BMJ Open. 1 juin 2020;10(10):e036814.

24. Hedible GB, Louart S, Neboua D, Catala L, Anago G, Sawadogo AG, et al. Evaluation of the routine implementation of pulse oximeters into integrated management of childhood illness (IMCI) guidelines at primary health care level in West Africa: the AIRE mixed-methods research protocol. BMC Health Serv Res. 24 déc 2022;22:1579.

25. Hedible GB, Sawadogo AG, Zair Z, Kargougou GD, Agbeci H, Meda B, et al. Identification of severe cases with routine Pulse Oximetry use into the Integrated Management of Childhood Illness at Primary Health Centres level in West Africa: A cross-sectional study within the AIRE project in Burkina Faso, Guinea, Mali and Niger, 2021 - 2022 [Internet]. medRxiv; 2024 [cité 16 oct 2024]. p. 2024.10.14.24315439. Disponible sur: https://www.medrxiv.org/content/10.1101/2024.10.14.24315439v1

26. Mémento de soins hospitaliers pédiatriques, OMS, 2ème édition, 2015 [Internet]. [cité 5 janv 2024]. Disponible sur: https://iris.who.int/bitstream/handle/10665/187940/9789242548372_fre.pdf?sequence=1

27. Duke T, Blaschke AJ, Sialis S, Bonkowsky JL. Hypoxaemia in acute respiratory and non-respiratory illnesses in neonates and children in a developing country. Arch Dis Child. 1 févr 2002;86(86):108–12.

28. McCollum ED, Nambiar B, Deula R, Zadutsa B, Bondo A, King C, et al. Impact of the 13-Valent Pneumococcal Conjugate Vaccine on Clinical and Hypoxemic Childhood Pneumonia over Three Years in Central Malawi: An Observational Study. PLOS ONE. 4 janv 2017;12(12):e0168209.

29. Louart S, Hedible GB, Balde H, Coulibaly A, Dagobi AE, Kadio K, et al. Acceptability of the routine use of pulse oximeters into the Integrated Management of Childhood Illness guidelines at primary health centers in West Africa: a mixed-methods study. [Internet]. medRxiv; 2024 [cité 18 oct 2024]. p. 2024.10.16.24315522. Disponible sur: https://www.medrxiv.org/content/10.1101/2024.10.16.24315522v1

30. Ginsburg AS, Tawiah Agyemang C, Ambler G, Delarosa J, Brunette W, Levari S, et al. mPneumonia, an Innovation for Diagnosing and Treating Childhood Pneumonia in Low-Resource Settings: A Feasibility, Usability and Acceptability Study in Ghana. PLoS ONE. 27 oct 2016;11(10):e0165201.

31. King C, Boyd N, Walker I, Zadutsa B, Baqui AH, Ahmed S, et al. Opportunities and barriers in paediatric pulse oximetry for pneumonia in low-resource clinical settings: a qualitative evaluation from Malawi and Bangladesh. BMJ Open. 30 janv 2018;8(8):e019177.

32. Rahman AE, Ameen S, Hossain AT, Perkins J, Jabeen S, Majid T, et al. Introducing pulse oximetry for outpatient management of childhood pneumonia: An implementation research adopting a district implementation model in selected rural facilities in Bangladesh. eClinicalMedicine [Internet]. 1 août 2022 [cité 23 juill 2023];50. Disponible sur: https://www.thelancet.com/journals/eclinm/article/PIIS2589-5370(22)00241-3/fulltext

33. Hedible GB, al. What are the challenges and needs before implementing routine pulse oximetry in IMCI consultations at primary health centers in West Africa? Baseline site assessment of the operational AIRE project. To be submitted to BMJ Global Health; 2024.

34. Beyene H, Kassa DH, Tadele H, Persson L, Defar A, Berhanu D. Factors associated with the referral of children with severe illnesses at primary care level in Ethiopia: a cross-sectional study. BMJ Open. 1 juin 2021;11(11):e047640.

35. Blanc J, Locatelli I, Rarau P, Mueller I, Genton B, Boillat-Blanco N, et al. Retrospective study on the usefulness of pulse oximetry for the identification of young children with severe illnesses and severe pneumonia in a rural outpatient clinic of Papua New Guinea. PLOS ONE. 15 avr 2019;14(14):e0213937.

36. Agbeci H, al. Direct and indirect household costs of care of children under 5 years old attending Integrated Management of Childhood Illness consultations at Primary Healthcare Centres in Burkina Faso, Guinea, Mali and Niger: a cross-sectional costing study nested in the longitudinal AIRE project 2021-2022. To be submitted to BMJ Global Health; 2024.

37. McCollum ED, Ahmed S, Roy AD, Islam AA, Schuh HB, King C, et al. Risk and accuracy of outpatient-identified hypoxaemia for death among suspected child pneumonia cases in rural Bangladesh: a multifacility prospective cohort study. Lancet Respir Med [Internet]. 7 avr 2023 [cité 18 juill 2023];0(0). Disponible sur: https://www.thelancet.com/journals/lanres/article/PIIS2213-2600(23)00098-X/fulltext

38. Bakare AA, Salako J, King C, Olojede OE, Bakare D, Olasupo O, et al. ‘Let him die in peace’: understanding caregiver’s refusal of medical oxygen treatment for children in Nigeria. BMJ Glob Health. 16 mai 2024;9(9):e014902.

39. Källander K, Counihan H, Cerveau T, Mbofana F. Barriers on the pathway to survival for children dying from treatable illnesses in Inhambane province, Mozambique. J Glob Health. juin 2019;9(1):010809.

40. Treleaven E, Whidden C, Cole F, Kayentao K, Traoré MB, Diakité D, et al. Relationship between symptoms, barriers to care and healthcare utilisation among children under five in rural Mali. Trop Med Int Health. août 2021;26(8):943–52.

41. Beyene H, Hailu D, Tadele H, Persson LÅ, Berhanu D. Insufficient referral practices of sick children in Ethiopia shown in a cross-sectional survey. Acta Paediatr Oslo Nor 1992. sept 2020;109(109):1867–74.

42. Moonpanane K, Pitchalard K, Thepsaw J, Singkhorn O, Potjanamart C. Healthcare service utilization of hill tribe children in underserved communities in thailand: Barriers to access. BMC Health Serv Res. 2 sept 2022;22:1114.

43. Evans MV, Andréambeloson T, Randriamihaja M, Ihantamalala F, Cordier L, Cowley G, et al. Geographic barriers to care persist at the community healthcare level: Evidence from rural Madagascar. PLOS Glob Public Health. 27 déc 2022;2(2):e0001028.

44. Jeffreys M, Smiler K, Ellison Loschmann L, Pledger M, Kennedy J, Cumming J. Consequences of barriers to primary health care for children in Aotearoa New Zealand. SSM - Popul Health. 5 févr 2022;17:101044.

45. McCollum ED, Bjornstad E, Preidis GA, Hosseinipour MC, Lufesi N. Multicenter study of hypoxemia prevalence and quality of oxygen treatment for hospitalized Malawian children. Trans R Soc Trop Med Hyg. mai 2013;107(5):285–92.

46. Duke T, Wandi F, Jonathan M, Matai S, Kaupa M, Saavu M, et al. Improved oxygen systems for childhood pneumonia: a multihospital effectiveness study in Papua New Guinea. The Lancet. 11 oct 2008;372(9646):1328–33.

47. Graham HR, Olojede OE, Bakare AA, Iuliano A, Olatunde O, Isah A, et al. Measuring oxygen access: lessons from health facility assessments in Lagos, Nigeria. BMJ Glob Health. 3 août 2021;6(6):e006069.

48. Hill SE, Njie O, Sanneh M, Jallow M, Peel D, Njie M, et al. Oxygen for treatment of severe pneumonia in The Gambia, West Africa: a situational analysis. Int J Tuberc Lung Dis. 1 mai 2009;13(13):587–93.

49. Baker T, Schell CO, Petersen DB, Sawe H, Khalid K, Mndolo S, et al. Essential care of critical illness must not be forgotten in the COVID-19 pandemic. Lancet Lond Engl. 2020;395(10232):1253–4.

50. Mangipudi S, Leather A, Seedat A, Davies J. Oxygen availability in sub-Saharan African countries: a call for data to inform service delivery. Lancet Glob Health. sept 2020;8(9):e1123–4.

51. 012_WHO-AFRO_Strategic-Response-to-COVID-19_A4_P_V3. 2021 [Internet]. [cité 9 sept 2023]. Disponible sur: https://www.afro.who.int/sites/default/files/2021-04/012_WHO-AFRO_Strategic-Response-to-COVID-19_A4_P_V3.indd%20-%20FINAL%20-%20FR%20FINAL.pdf

52. Duke T, Graham SM, Cherian MN, Ginsburg AS, English M, Howie S, et al. Oxygen is an essential medicine: a call for international action. Int J Tuberc Lung Dis Off J Int Union Tuberc Lung Dis. nov 2010;14(11):1362–8.

53. Graham HR, Kitutu FE, Kamuntu Y, Kunihira B, Engol S, Miller J, et al. Improving effective coverage of medical-oxygen services for neonates and children in health facilities in Uganda: a before–after intervention study. Lancet Glob Health. 14 août 2024;12(12):e1506–16.

54. Budu E, Seidu AA, Ameyaw EK, Agbaglo E, Adu C, Commey F, et al. Factors associated with healthcare seeking for childhood illnesses among mothers of children under five in Chad. PLoS ONE. 5 août 2021;16(16):e0254885.

55. Cassy A, Chicumbe S, Saifodine A, Zulliger R. Factors associated with malaria care seeking among children under 5 years of age in Mozambique: a secondary analysis of the 2018 Malaria Indicator Survey. Malar J. 24 mars 2022;21:100.

56. Rees CP, Hawkesworth S, Moore SE, Dondeh BL, Unger SA. Factors Affecting Access to Healthcare: An Observational Study of Children under 5 Years of Age Presenting to a Rural Gambian Primary Healthcare Centre. PLoS ONE. 23 juin 2016;11(11):e0157790.

57. Hooli S, King C, McCollum ED, Colbourn T, Lufesi N, Mwansambo C, et al. In-hospital mortality risk stratification in children aged under 5 years with pneumonia with or without pulse oximetry: A secondary analysis of the Pneumonia REsearch Partnership to Assess WHO REcommendations (PREPARE) dataset. Int J Infect Dis. 1 avr 2023;129:240–50.

58. Wilkes C, Bava M, Graham HR, Duke T. What are the risk factors for death among children with pneumonia in low- and middle-income countries? A systematic review. J Glob Health. 13:05003.

59. Graham HR, King C, Duke T, Ahmed S, Baqui AH, Colbourn T, et al. Hypoxaemia and risk of death among children: rethinking oxygen saturation, risk-stratification, and the role of pulse oximetry in primary care. Lancet Glob Health. 1 août 2024;12(8):e1359–64.

